# Buried or Exposed Kirschner Wire for the Management of Hand and Forearm Fractures: A Systematic Review, Meta-Analysis, and Meta-Regression

**DOI:** 10.1101/2023.12.07.23299683

**Authors:** Nucki Nursjamsi Hidajat, RM. Satrio Nugroho Magetsari, Gregorius Thomas Prasetiyo, Danendra Rakha Putra Respati, Kevin Christian Tjandra

## Abstract

**Background:** The recommendation on whether to bury or expose the Kirschner wire (K-wire) for the management of fractures has still been controversial with inconsistent results in the published studies. This study aims to summarize the comparison between buried and exposed K-wire for the management of hand and forearm fractures.

**Methods:** We conducted relevant literature searches on Europe PMC, Medline, Scopus, and Cochrane Library databases using specific keywords. The results of continuous variables were pooled into the standardized mean difference (SMD), while dichotomous variables were pooled into odds ratio (OR) along with 95% confidence intervals (95% CI) using random-effect models.

**Results:** A total of 11 studies were included. Our pooled analysis revealed that buried K-wire was associated with a lower risk of pin site infection [RR 0.49 (95% CI 0.36 – 0.67), *p* < 0.00001, *I*^2^ = 0%] and longer duration until pin removal [MD 33.85 days (95% CI 18.68 – 49.02), *p* < 0.0001, *I*^2^ = 99%] when compared with exposed K-wire. However, the duration of surgery was significantly longer in the buried K-wire [MD 6.98 minutes (95% CI 2.19 – 11.76), *p* = 0.004, *I*^2^ = 42%] with no significant difference in the early pin removal rate [RR 0.73 (95% CI 0.36 – 1.45), *p* = 0.37, *I*^2^ = 0%]. Further regression analysis revealed that sample size, age, sex, and duration of follow-up did not affect those relationships.

**Conclusion:** Buried K-wire may offer benefits in reducing the infection rate with a longer duration until pin removal. However, further RCTs with larger sample sizes are still needed to confirm the results of our study.

## INTRODUCTION

A fracture is defined as a break in the continuity of the bone tissue structure, which can be caused by trauma or non-trauma.[1] Forearm fractures involving the radius and ulna were the most common fracture of the upper limb with an annual incidence of 16.2 fractures per 10,000 individuals, followed by hand fractures involving the metacarpals and phalanx with an annual incidence of 12.5 and 6.4 fractures per 10,000 individuals, respectively.[2] Both forearms and hands are essential in carrying out daily activities so these two types of fractures often result in the disruption of a person’s quality of life.[2]

Kirschner wire (K-wire) is often used by orthopedic surgeons, especially those specializing in the field of hand and upper extremity surgery to provide fixation for unstable hand or forearm fractures because it is associated with good outcomes at a relatively low cost.[3,4] Results from several previous studies have demonstrated the non-inferiority of K-wire when compared to plate-screw fixation in the management of unstable metacarpal and phalangeal fractures.[5,6] A meta-analysis study involving 5 randomized clinical trials (RCTs) and 9 cohort studies showed similar clinical, functional, and radiological outcomes between percutaneous K-wires when compared with volar locking plates (VLPs) for distal radius fixation, but with lower re-operation risk in the K-wire group.[7] These wires can be implanted under the skin or left exposed. A national survey conducted in the United Kingdom (UK) showed that the majority of orthopedic surgeons, plastic surgeons, and junior surgical trainees chose to leave K-wire not buried because it was easier to take.[8] The concern that may arise from exposed K-wire is the potentially greater risk of infection because it has direct contact with the outside air and environmental exposure.[8]

Several studies have been conducted to compare buried and exposed K-wires in the management of hand and forearm fractures but have yielded inconsistent results.[9,10] A randomized trial study conducted by Maradei-Pereira JAR et al.[9] distal radial, fractures showed a greater risk of infection when K-wires were left exposed than when they were buried. On the other hand, Khaled M et al.[10] showed no significant difference in the complication rate, including pin infection incidence between exposed and buried K-wire. Given these inconsistencies, a meta-analysis may help. This systematic review and meta-analysis aim to summarize the latest evidence regarding the comparison between exposed and buried K-wires for the management of hand and forearm fractures.

## Material and Method

### Registration

The Preferred Reporting Items for Systematic Reviews and Meta-Analyses (PRISMA) were used for this project. On December 4^th^, 2023, this systematic review and meta-analysis was registered to the Open Science Framework (OSF). The registration was identified as Buried or Exposed Kirschner Wire for the Management of Hand and Forearm Fractures: A Systematic Review, Meta-Analysis, and Meta-Regression. https://doi.org/10.17605/OSF.IO/647WF

### Eligibility Criteria

This review was written following the PRISMA statement and Cochrane Handbook guidelines.[11,12] We included the following studies: (1) studies on patients of any age with the diagnosis of hand (phalangeal or metacarpal) or forearm (radius or ulna) fractures who have undergone surgery with Kirschner wire (K-wire) fixation (Population); (2) compare between buried and exposed K-wire fixation (Intervention and Control); (3) have data on the primary outcome (pin infection) with/without secondary outcomes (early pin removal, days to pin removal, and duration of surgery) (Outcome); (4) presented in the form of observational studies (cohort/case-control) or randomized clinical trials (RCTs) (Study Design).

Meanwhile, the studies (1) conducted in patients with humeral, shoulder, or lower extremity fractures; (2) using another method of fixation besides K-wire for the patients; (3) presented in the form of case reports, case series, and review articles were excluded from our analysis.

### Literature Search and Study Selection

The search of English literature on 4 international databases: Medline, Scopus, Europe PMC, and Cochrane Library was performed by five independent authors from the date of inception until March 7^th^, 2023. We used the following combined keywords to capture all potentially eligible literature: “(buried OR implanted OR concealed OR embedded) AND (exposed OR uncovered) AND (Kirschner wire OR K-wire) AND (hand fractures OR phalangeal fractures OR metacarpal fractures OR forearm fractures OR radius fractures OR ulna fractures)” as shown in **Table 1**. At first, the identified articles from those databases were removed for duplicates and then screened based on their titles/abstracts. These articles that passed the initial screening process underwent the second step of evaluation in the form of full-text to see their compatibility with our inclusion/exclusion criteria. All of these processes for article selection were conducted by the same five authors who performed the literature searching process. All discrepancies were resolved through discussion with the third author.

**Table 1.** Literature search strategy.

### Data Extraction and Quality Assessment

We extracted the following data for analytical purposes: study ID, publication year, country, study design, sample size, baseline characteristics of participants (mean age and sex distribution), follow-up duration, the number of participants in the buried and exposed K-wire groups, and the outcomes of interest. These data extracted by five independent authors were tabulated into Microsoft Excel 2019.

The outcomes of interest in this study were divided into primary and secondary outcomes. The primary outcome was pin site infection which encompassed both superficial and deep infection at or around the site of K-wire placement. The secondary outcomes consisted of early pin removal rate, days to pin removal, and duration of surgery. Early pin removal was defined as the removal of K-wire before the initially planned date due to complications.

### Risk of Bias Assessment

The risk of bias assessment was conducted by five independent authors using validated tools. For analyzing the quality of included RCTs, we used a tool from Cochrane Collaborations, namely Risk of Bias version 2 (RoB v2), which includes a methodological assessment of 5 domains: (a) randomization process; (b) deviations from intended interventions; (c) missing outcome data; (d) measurement of the outcome; and (e) selection of the reported results.[13] The result is shown in **Fig 2**. The authors’ evaluations were categorized as “low risk,” “high risk,” or “some concerns” of bias.[13] For analyzing the quality of included cohort/case-control studies, we used Newcastle-Ottawa Scale (NOS) from the Ottawa Hospital Research Institute (OHRI) which includes 3 domains of assessment: (1) selection of participants; (2) comparability between exposed and non-exposed cohort; and (3) outcome ascertainment.[14] The total score that can be achieved ranged from 0 to 9 where articles with ≥7 scores were considered as having “good” qualities.[14]

### Statistical Analysis

We used mean difference (SMD) along with 95% confidence intervals (95% CI) for the analytical pooling of continuous variables outcomes by using the Inverse-Variance formula. We also pooled dichotomous variable outcomes into risk ratio (RR) along with 95% CI by using the Mantel-Haenszel formula. Random-effect models were chosen in this review because of the consideration that significant heterogeneity was expected due to differences in the population characteristics and differences in the duration of follow-up. In this review, we used the I-squared (I^2^) statistic to assess the heterogeneity between studies where I^2^ values of >50% were categorized as significant heterogeneity. We used a combined formula from Luo D et al.[15] and Wan X et al.[16] to change the data expressed in the form of the median and interquartile range (IQR) or data expressed as median, minimum, and maximum into mean and standard deviations (SD) for pooled analysis purposes. A publication bias analysis was performed when there were more than 10 studies on each outcome of interest. All of these statistical analyses were carried out using an application from the Cochrane Collaboration, namely Review Manager 5.4.

## RESULTS

### Study Selection and Characteristics

A literature search on 4 international databases yielded a total of 166 studies. After removing duplicates and screening studies based on their titles and abstracts, 140 studies were removed, leaving 26 studies. These 26 studies were assessed in a full-text form where 15 studies did not meet our eligibility criteria as follows: 8 studies were conducted on patients with humeral fractures, 2 studies did not have any control group, 2 studies did not have data on the outcome of interest, 1 study was only review article, 1 study did not use K-wire for all of the participants, and 1 study was not published in the English language, thus only 11 studies[9,10,17–25] were included in the final analysis **Fig 1**. Of these 11 studies, 5 were prospective RCTs and 6 were retrospective cohort studies. The number of samples ranged from 52 to 695 with the duration of follow-up varying from 6 weeks to 1 year. A summary of the baseline characteristics of the included studies can be found in **Table 2**.

**Figure 1.** PRISMA diagram of the detailed process of selection of studies for inclusion in the systematic review and meta-analysis.

**Table 2.** Characteristics of included studies.

### Quality of Study Assessment

The risk of bias assessment by using the RoB v2 tool revealed that only 1 RCT[9] had a “low risk” of bias in all five assessment domains. The remaining four RCTs[10,17–19] were judged to have “some concern” risk of bias. One RCT[18] used consecutive non-random sampling for the selection of participants which was deemed an inappropriate method for randomization, but there was no significant difference in the baseline characteristics of participants in the two groups of intervention, suggesting no serious problems during randomization, therefore was judged to have “some concern” risk of bias in the randomization process. Two RCTs[10,17] did not mention in detail the blinding of the outcome assessors, therefore were judged to have “some concern” risk of bias in the measurement of the outcome. Lastly, one remaining RCT[19] did not mention in detail regarding both the randomization method and “blinding” of the outcome assessors, therefore were also judged to have “some concern” risk of bias in the randomization process and measurement of the outcome. The evaluation of all included cohort studies by using the NOS tool revealed “good quality” studies with total scores ranging from 7 to 8. The summary of the risk of bias assessment for all included studies in this review can be found in **Fig 2** and **Table 3**.

**Table 3.** Newcastle-Ottawa quality assessment of observational studies.

### Primary Outcome

#### Pin Site Infection

Based on our pooled analysis of 11 studies (n = 2,022), it has been shown that buried K-wire was associated with a lower risk of pin site infection when compared with exposed K-wire in patients with hand and forearm fractures [RR 0.49 (95% CI 0.36 – 0.67), *p* < 0.00001, *I*^2^ = 0%, random-effect models] as shown in **Fig 3**. Subgroup analysis based on the study design revealed consistent and significant results for both RCTs [RR 0.38 (95% CI 0.21 – 0.68), *p* = 0.001, *I*^2^ = 0%, random-effect models] and observational studies [RR 0.54 (95% CI 0.37 – 0.79), *p* = 0.001, *I*^2^ = 0%, random-effect models] with a lower RR was found in the RCTs studies.

**Figure 3.** Forest plot that demonstrates the comparison between buried and exposed K-wire for the pin site infection outcome in both randomized clinical trials (RCTs) and observational studies.

### Secondary Outcome

#### Early Pin Removal

Pooled analysis from 6 studies (n = 1,200) showed a non-significant difference in the rate of early pin removal between buried and exposed K-wire for patients with hand and forearm fractures [RR 0.73 (95% CI 0.36 – 1.45), *p* = 0.37, *I*^2^ = 0%, random-effect models] as in **Fig 4**. Subgroup analysis based on the study design revealed that the results remained non-significant for both RCT [RR 0.65 (95% CI 0.10 – 4.03), *p* = 0.64, *I*^2^ = 31%, random-effect models] and observational studies [RR 0.74 (95% CI 0.34 – 1.61), *p* = 0.44, *I*^2^ = 0%, random-effect models].

**Figure 4.** Forest plot that demonstrates the comparison between buried and exposed K-wire for the early pin removal outcome in both randomized clinical trials (RCTs) and observational studies.

#### Days to Pin Removal

Pooled analysis from 4 studies (n = 1,493) showed that buried K-wire was associated with significantly longer days to pin removal when compared to exposed K-wire in patients with hand and forearm fractures [MD 33.85 days (95% CI 18.68 – 49.02), *p* < 0.0001, *I*^2^ = 99%, random-effect models]. The forest plot is available in **Fig 5** Subgroup analysis based on the study design revealed significant results for both RCT [MD 29.00 (95% CI 24.04 – 33.96), *p* < 0.00001, random-effect models] and observational studies [MD 32.53 (95% CI 16.89 – 48.18), *p* < 0.0001, *I*^2^ = 99%, random-effect models] with the MD was found to be higher in the observational studies.

**Figure 5.** Forest plot that demonstrates the comparison between buried and exposed K-wire for the days to pin removal outcome in both randomized clinical trials (RCTs) and observational studies.

#### Duration of Surgery

Pooled analysis from 3 studies (n = 325) showed that the duration of surgery was significantly longer for the buried K-wire when compared to exposed K-wire in patients with hand and forearm fractures [MD 6.98 minutes (95% CI 2.19 – 11.76), *p* = 0.004, *I*^2^ = 42%, random-effect models] as shown in **Fig 6**. Subgroup analysis based on the study design revealed that a significant difference in the duration of surgery was only seen in the RCTs studies [MD 7.53 (95% CI 0.74 – 14.31), *p* = 0.03, *I^2^* = 70%, random-effect models] but not in the observational studies [MD 5.45 minutes (95% CI −5.14 – 16.04), *p* = 0.31, random-effect models].

**Figure 6.** Forest plot that demonstrates the comparison between buried and exposed K-wire for the duration of surgery outcome in both randomized clinical trials (RCTs) and observational studies.

#### Meta-Regression

Identification of risk factors that influence the relationship of buried and exposed K-wire with all of the outcomes of interest was done with meta-regression. Our meta-regression revealed that variability in those outcomes in hand and forearm fractures patients receiving buried or exposed K-wire fixation cannot be explained by known patient factors associated with predictors of treatment outcomes (Table 4). From our meta-regression analysis, it was revealed that pin site infection in hand or forearm fractures patients treated with either buried or exposed K-wire was not significantly influenced by sample size (*p* = 0.3469) (**Fig 7A**), age (*p* = 0.5300) (**Fig 7B**), sex (*p* = 0.1105) (**Fig 7C**), or follow-up duration (*p* = 0.2895) (**Fig 7D**).

**Figure 7.** Bubble-plot for meta-regression. Meta-regression analysis showed that the comparison between buried and exposed K-wire with the pin site infection outcome was not significantly affected by sample size (A), age (B), sex (C), nor follow-up duration (D)

**Table 4.** Result for the meta-regression models for each outcome of interest.

The association between either buried or exposed K-wire with early pin removal was not significantly influenced by sample size (*p* = 0.8827) (**Fig 8A**), age (*p* = 0.4780) (**Fig 8B**), sex (*p* = 0.5103) (**Fig 8C**), or follow-up duration (*p* = 0.2063) (**Fig 8D**).

**Figure 8.** Bubble-plot for meta-regression. Meta-regression analysis showed that the comparison between buried and exposed K-wire with the early pin removal outcome was not significantly affected by sample size (A), age (B), sex (C), nor follow-up duration (D)

Our meta-regression analysis also revealed that the days to pin removal in patients with hand or forearm fractures receiving either buried or exposed K-wire were not significantly influenced by sample size (*p* = 0.1839) (**Fig 9A**), age (*p* = 0.0514) (**Fig 9B**), or sex (*p* = 0.8928) (**Fig 9C**). Variable “follow-up duration” was not possible to be included in the meta-regression analysis for this outcome because of the insufficient number of data in the included studies.

**Figure 9.** Bubble-plot for meta-regression. Meta-regression analysis showed that the comparison between buried and exposed K-wire with the days to pin removal outcome was not significantly affected by sample size (A), age (B), sex (C), nor follow-up duration (D).

Finally, the outcome “duration of surgery” was also not possible to be analyzed in the meta-regression analysis due to the insufficient number of included studies for this outcome.

All the meta-regression results for each outcome of interest is summarized in this below **Table 4**.

### Publication Bias

The Funnel-plot analysis shown in **Fig 10** for the pin site infection outcome revealed a relatively symmetrical inverted plot, suggesting no indication of publication bias. Meanwhile, the publication bias analysis was not performed for the early pin removal, days to pin removal, and duration of surgery outcomes due to less than 10 studies included in these outcomes where funnel plots and statistical tests to detect publication bias are less reliable.[26,27]

**Figure 10.** Funnel plot analysis that showed a relatively symmetrical inverted plot for the pin site infection outcome, indicating no publication bias.

## DISCUSSION

Based on the results of our meta-analysis, it has been shown that buried K-wire was associated with a lower risk of pin site infection and longer duration of pin removal when compared with exposed K-wire in patients with hand and forearm fractures, although the early pin removal rate did not differ significantly between two groups and the duration of surgery was significantly longer in the buried K-wire group. Further regression analysis also revealed that the study’s variables, such as sample size, age, sex, and follow-up duration did not significantly influence these relationships.

The results of our meta-analysis were in line with the results of the previous study by Chen L et al.[28] which also showed that buried K-wires had a lower risk of infection compared to exposed K-wires even though early pin removal did not differ significantly between groups. However, there were some differences between our current meta-analysis and the previous study by Chen L et al.[28]

First, the previous study by Chen L et al.[28] only included a total of 7 studies consisting of 2 RCTs and 5 observational studies. Meanwhile, our current meta-analysis has included more studies, a total of 11 studies (5 RCTs and 6 observational studies) with an additional 3 RCTs and 1 observational study when compared to the previous study which will certainly produce more solid evidence.

Second, the previous study by Chen L et al.[28] combined the results from RCT studies with results from observational studies. This action was inappropriate and not recommended when we referred to the guidelines from the Cochrane Handbook of Systematic Reviews of Intervention.[12] Observational studies tend to be susceptible to several biases such as selection bias and information bias which can have an impact on the research results.[29,30] Selection bias can lead to differences in the baseline characteristics of the two groups of participants which will also influence the results of the analysis.[29,30] Information bias can lead to inaccuracies of data and the results of outcome measurement obtained.[29,30] In addition, observational studies are often unable to anticipate the existence of several confounders which can also have an impact on research results.[29,30] Meanwhile, RCTs can avoid the presence of confounders through a process of randomizing the participants.[31,32] The existence of bias such as selection bias and information bias can also be minimized by allocation concealment and blinding methods, both for the participants and the outcome assessors.[31,32] Therefore, it is advisable to separate the results from RCTs and the results from observational studies. In our current meta-analysis, we have followed the recommendations of the Cochrane guidelines by separating the results from the RCTs and observational studies that can be seen in all of our forest plots, although there were no significant differences in results between those two study designs.

Third, our current meta-analysis had an additional outcome in the form of “days to pin removal” which was not analyzed in the previous study by Chen L et al.[28] From this new outcome, it was found that buried K-wire was able to provide a longer time for pin removal when compared to exposed K-wire. In addition, our current meta-analysis was also equipped with a meta-regression analysis to see the effect of several study variables, such as sample size, age, sex, and follow-up duration. From the results of our regression analysis, we have found no significant effect of these variables on the outcomes of interest.

Despite having several advantages above, our current meta-analysis is not without limitations. The majority of included studies have small sample sizes, under 100 participants. Some of these studies, especially the RCTs, also have “some concern” risk of bias caused by a lack of detailed information regarding the randomization method and the blinding of the outcome assessors. Two of our outcomes of interest, namely days to pin removal and duration of surgery, have relatively high heterogeneities (above 50%) which may be due to differences in the surgeon’s experience and length of follow-up duration. Finally, data regarding the cost-effectiveness analysis of the two K-wire methods were lacking in the included studies, therefore cannot be analyzed further. Further RCTs with larger sample sizes are still needed to confirm the results of our meta-analysis.

## CONCLUSION

Our systematic review dan meta-analysis suggests that buried K-wire may be more beneficial than exposed K-wire to provide a lower risk of infection and longer duration until pin removal in patients with hand and forearm fractures. However, the duration of surgery was relatively longer in the buried K-wire groups with no significant difference in the rate of early pin removal. The final decision on whether to bury or expose the K-wire fixation should still be based on the surgeon’s judgment with the consideration of the patient’s clinical condition as well as the benefits and risks for each patient. Further RCTs with larger sample sizes are still needed to confirm the results of our study.

## Data Availability

All relevant data are within the manuscript and its Supporting Information files.

## STATEMENTS AND DECLARATIONS

### Availability of data and materials

#### Underlying data

All data underlying the results are available as part of the article and no additional source data are required.

#### Reporting guidelines

Mendeley Data: Buried or Exposed Kirschner Wire for the Management of Hand and Forearm Fractures: A Systematic Review, Meta-Analysis, and Meta-Regression. doi: 10.17632/cxjd9nmx8r.1.

Data are available under the terms of the Creative Commons Attribution 4.0 Internationallicense (CC-BY 4.0).

## Acknowledgment

None

## Ethical Approval

Not applicable

## Patient’s Consent for Publication

Not applicable

## Supporting Information

**S1 Fig. Figure 1. PRISMA diagram of the detailed process of selection of studies for inclusion in the systematic review and meta-analysis.**

**S2 Fig. Figure 2. Risk of Bias assessment of the included studies using RoB v2 tool**

**S3 Fig. Figure 3. Forest plot that demonstrates the comparison between buried and exposed K-wire for the pin site infection outcome in both randomized clinical trials (RCTs) and observational studies.**

**S4 Fig. Figure 4. Forest plot that demonstrates the comparison between buried and exposed K-wire for the early pin removal outcome in both randomized clinical trials (RCTs) and observational studies.**

**S5 Fig. Figure 5. Forest plot that demonstrates the comparison between buried and exposed K-wire for the days to pin removal outcome in both randomized clinical trials (RCTs) and observational studies.**

**S6. Fig. Figure 6. Forest plot that demonstrates the comparison between buried and exposed K-wire for the duration of surgery outcome in both randomized clinical trials (RCTs) and observational studies.**

**S7. Fig. Figure 7. Bubble-plot for meta-regression. Meta-regression analysis showed that the comparison between buried and exposed K-wire with the pin site infection outcome was not significantly affected by sample size (A), age (B), sex (C), nor follow-up duration (D)**

**S8. Fig. Figure 8. Bubble-plot for meta-regression. Meta-regression analysis showed that the comparison between buried and exposed K-wire with the early pin removal outcome was not significantly affected by sample size (A), age (B), sex (C), nor follow-up duration (D)**

**S9. Fig. Figure 9. Bubble-plot for meta-regression. Meta-regression analysis showed that the comparison between buried and exposed K-wire with the days to pin removal outcome was not significantly affected by sample size (A), age (B), sex (C), nor follow-up duration (D).**

**S10 Fig. Figure 10. Funnel plot analysis that showed a relatively symmetrical inverted plot for the pin site infection outcome, indicating no publication bias.**

**S1 Table. Table 1. Literature search strategy**

**S2 Table. Table 2. Characteristics of included studies**

**S3 Table. Table 3. Newcastle-Ottawa quality assessment of observational studies**

**S4 table. Table 4. Result for the meta-regression models for each outcome of interest**

---

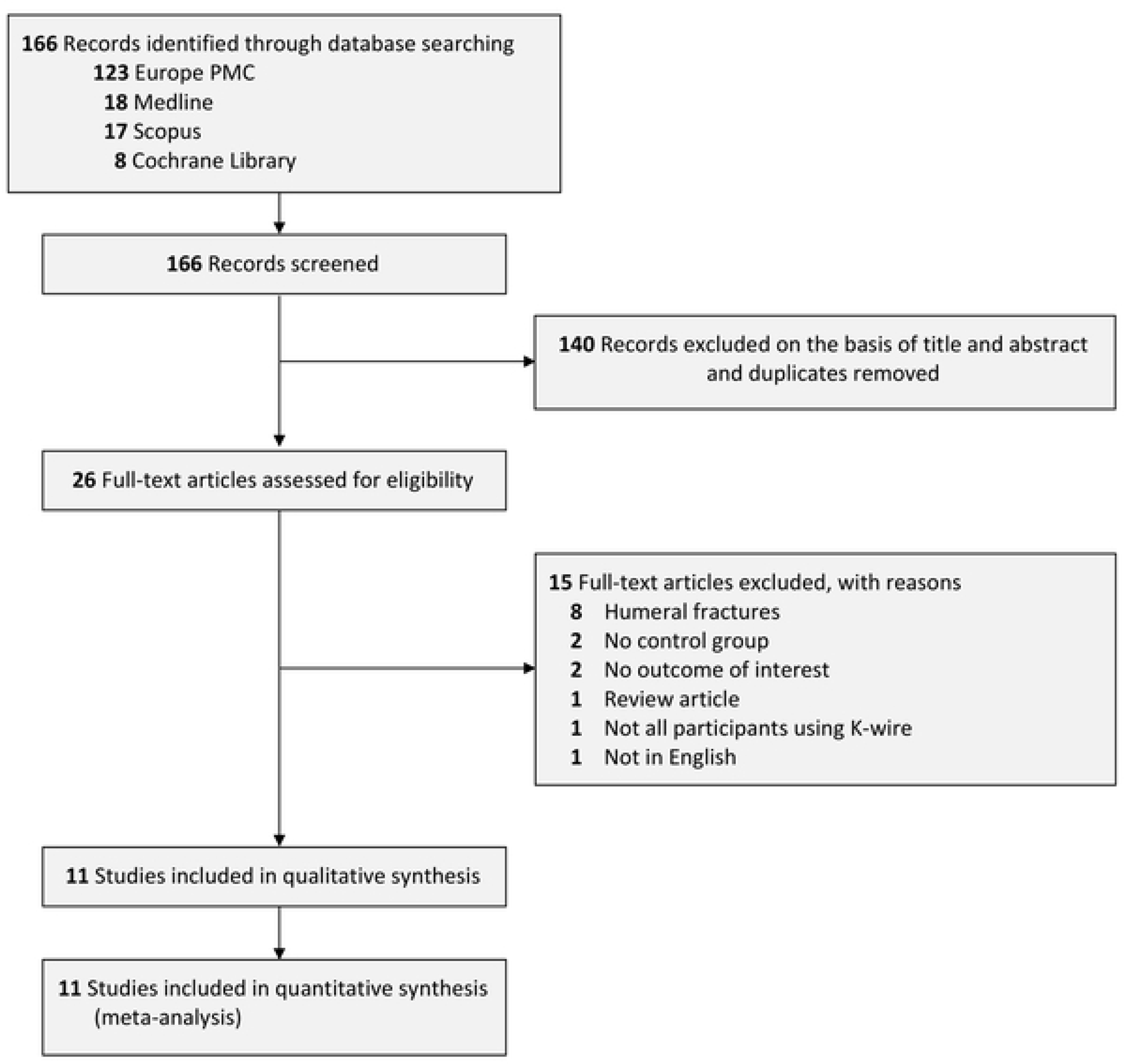

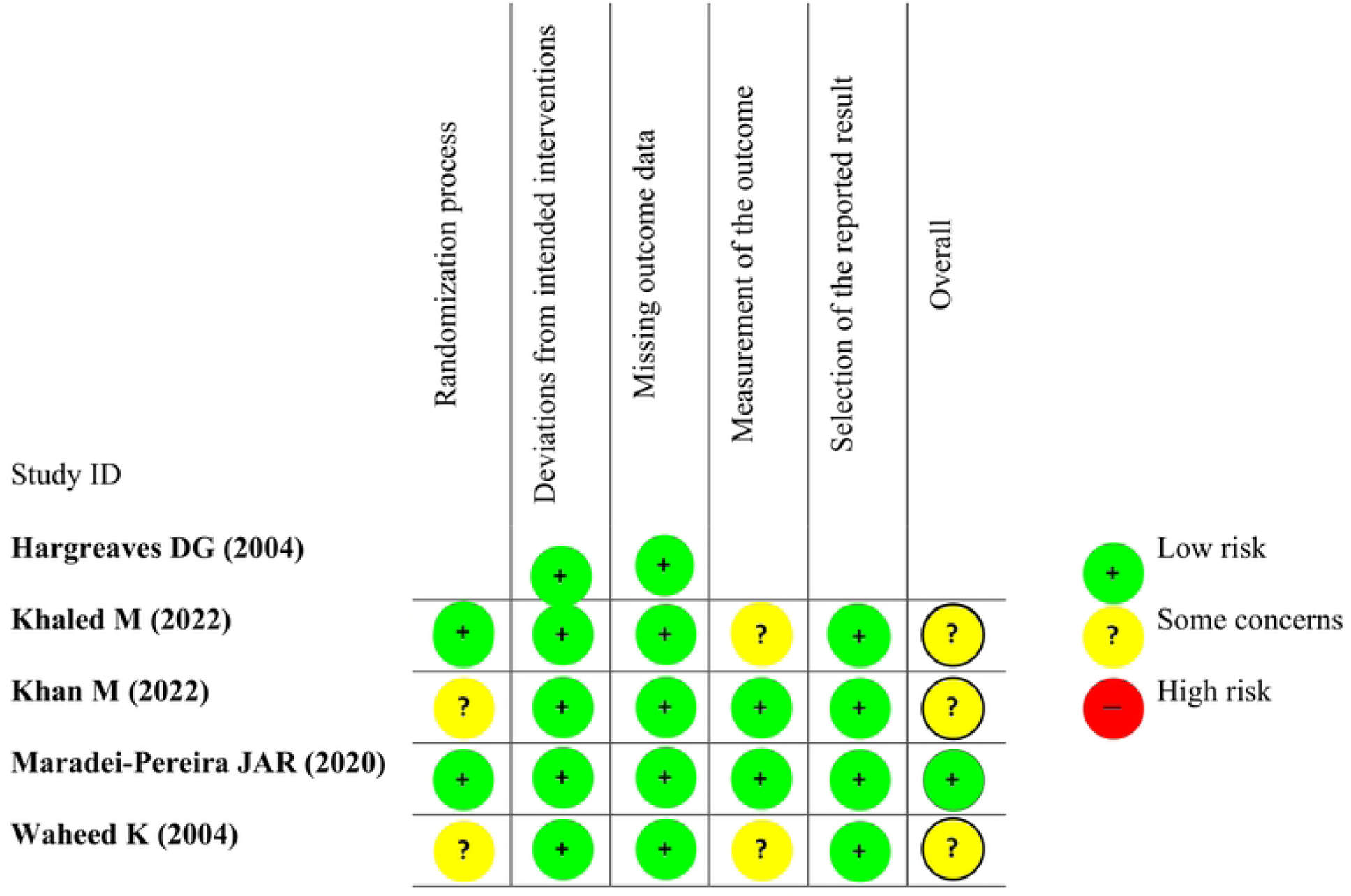

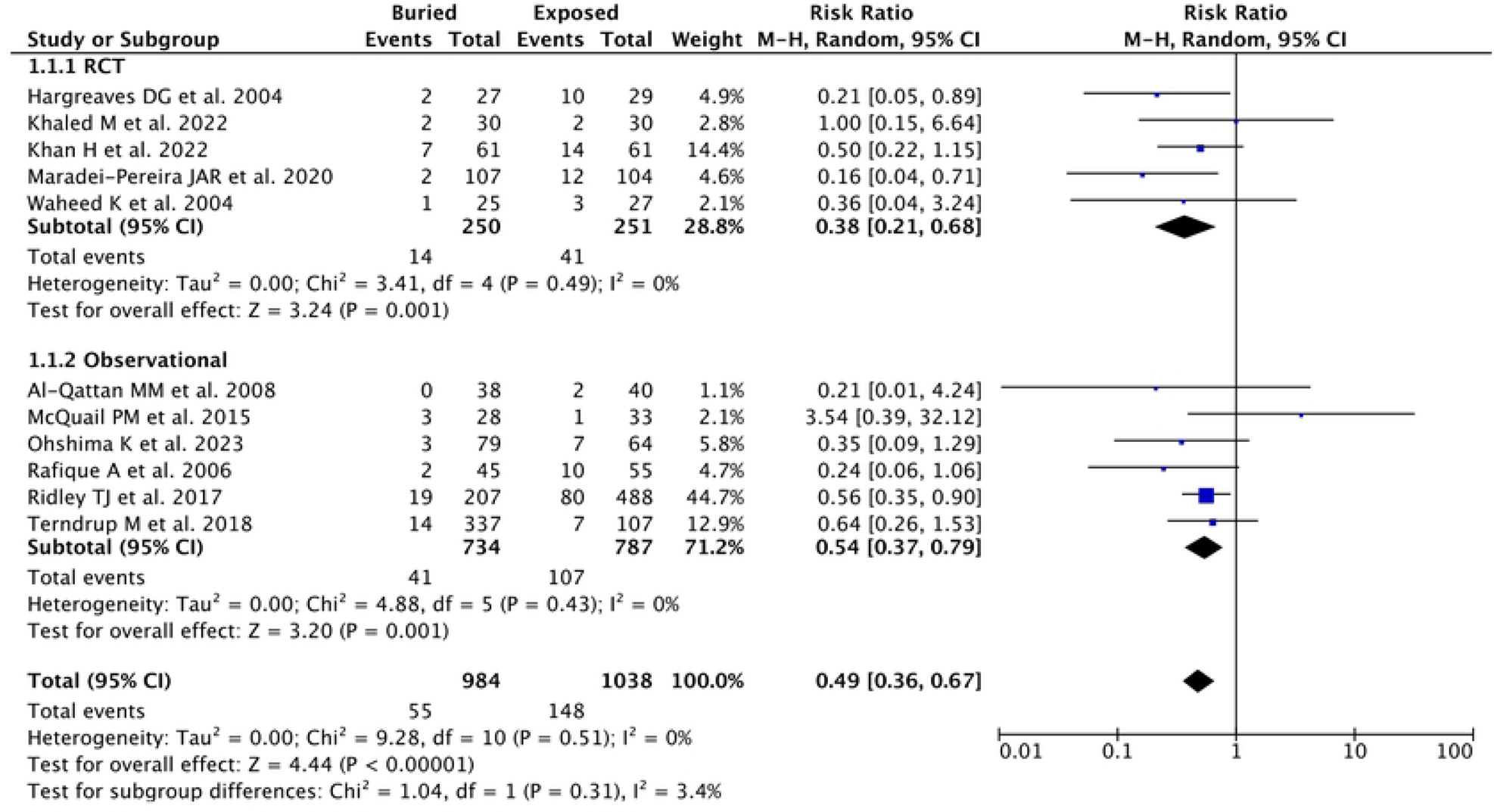

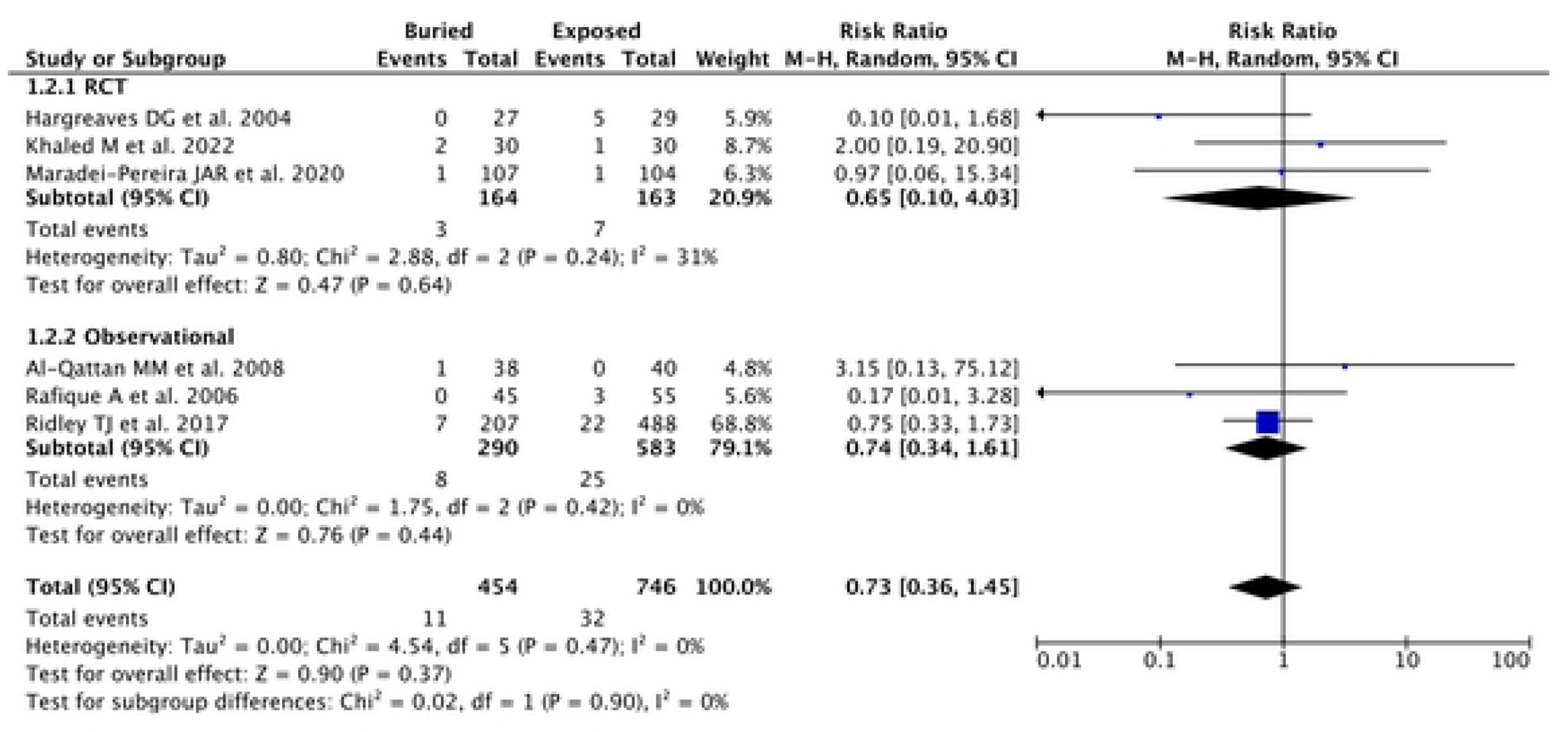

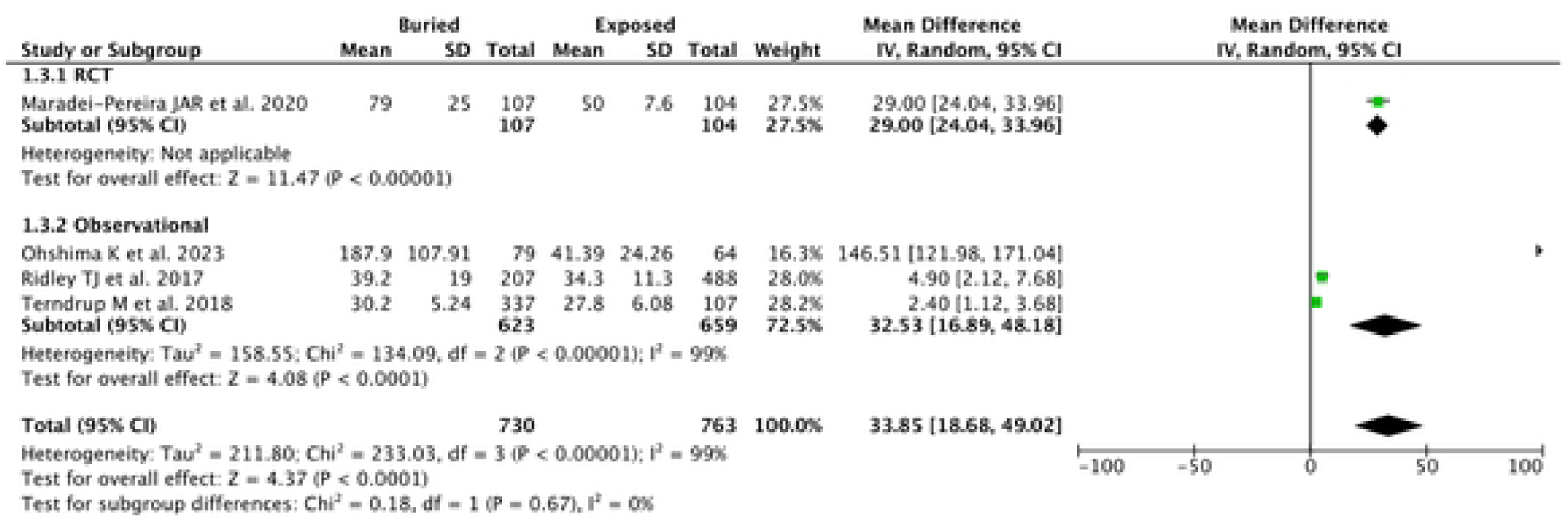

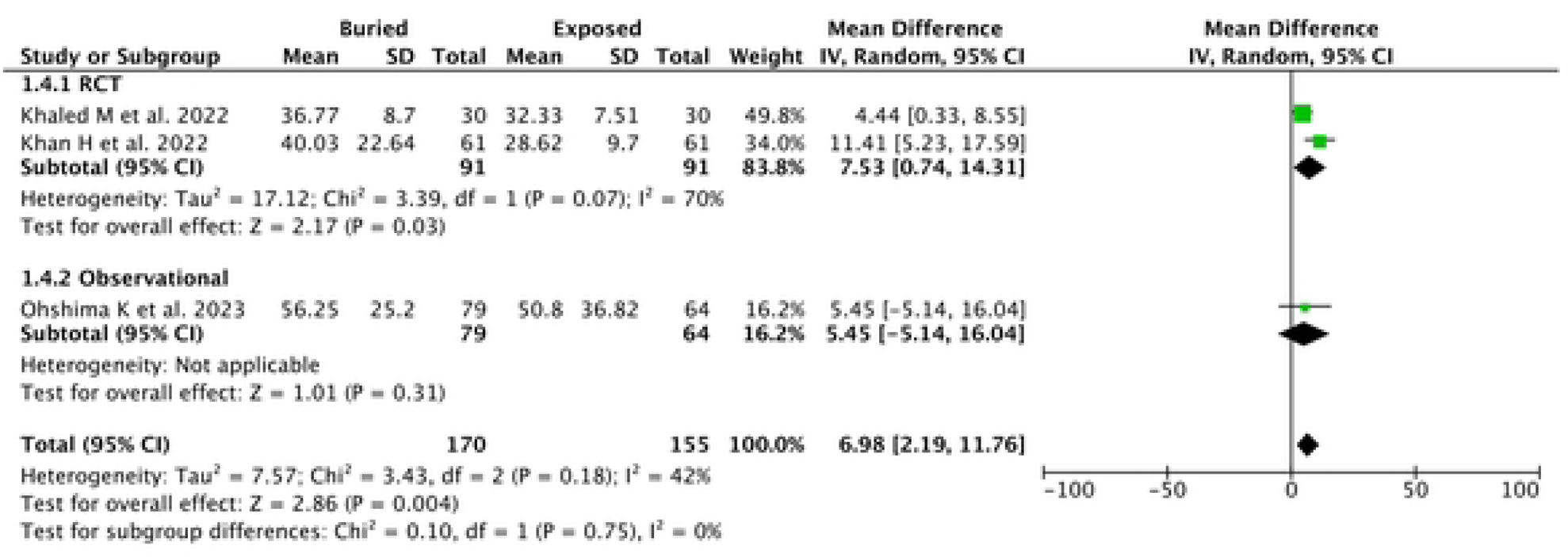

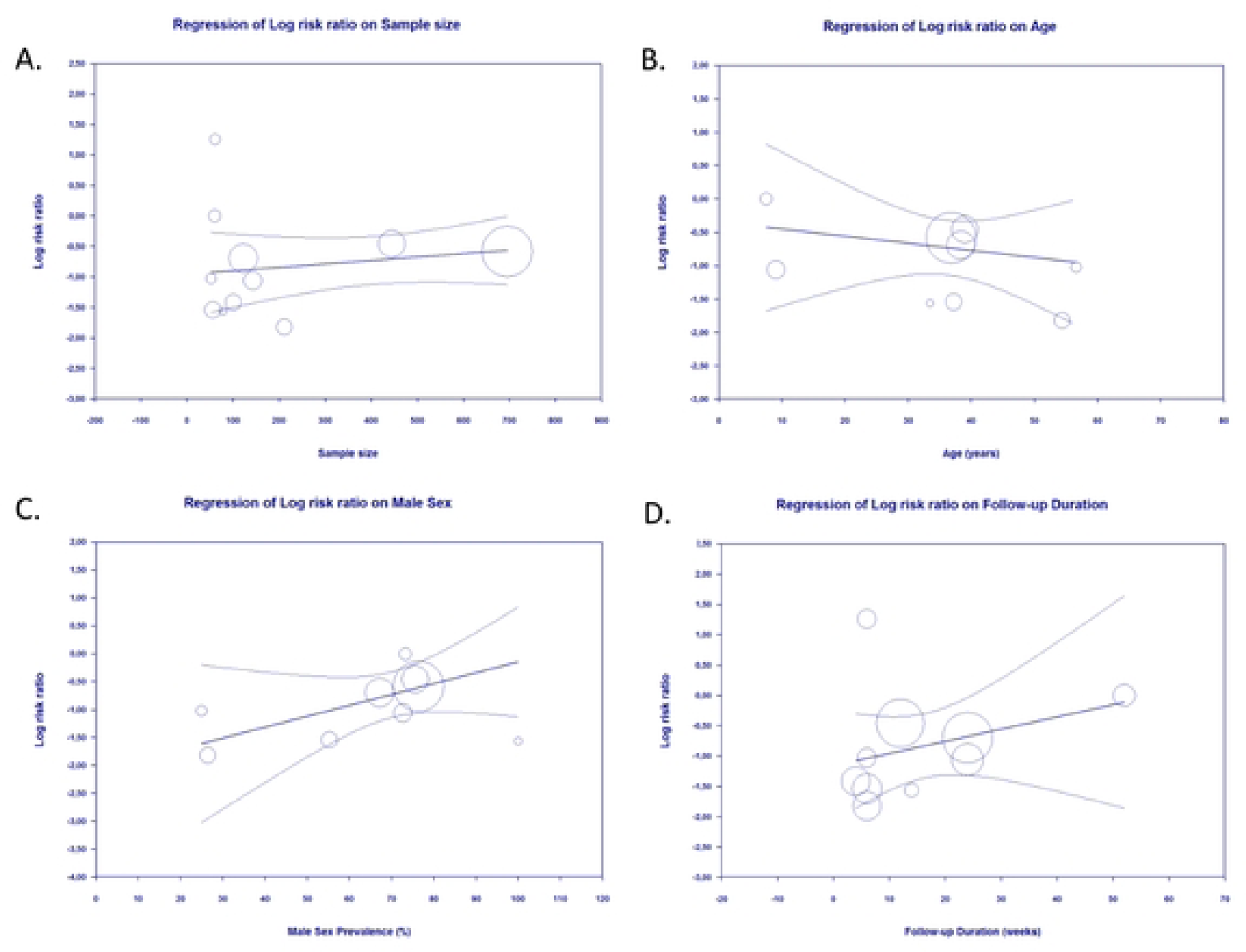

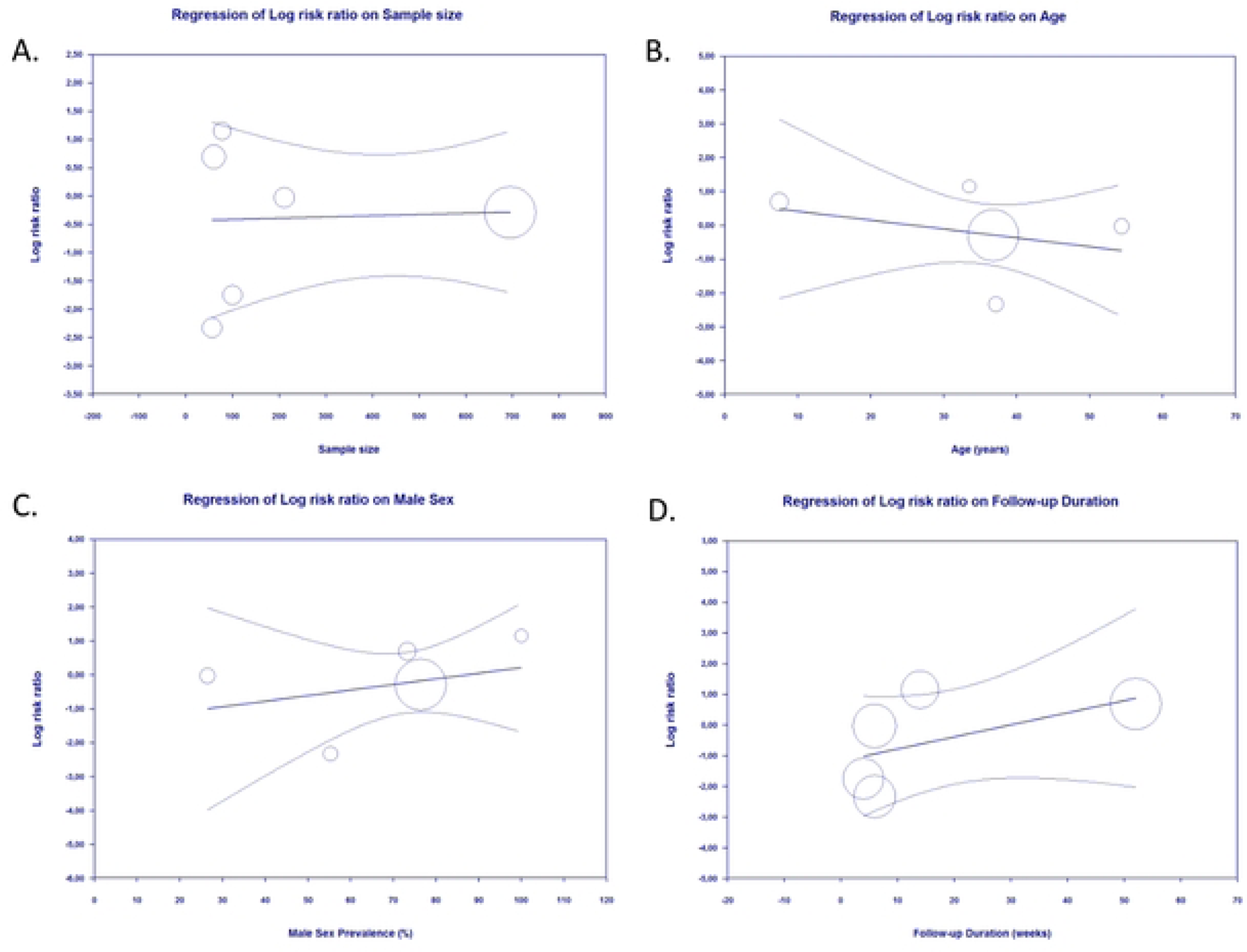

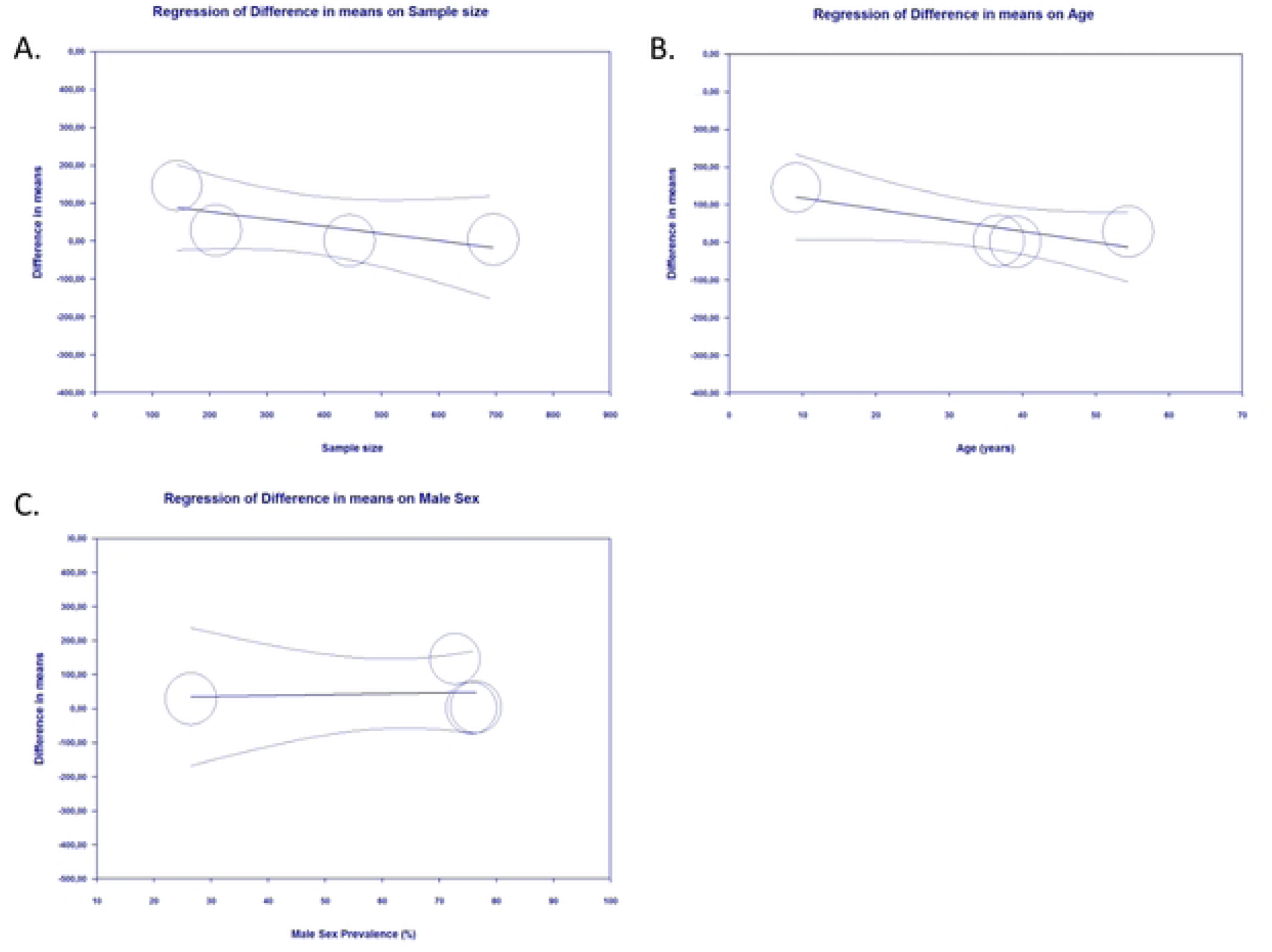

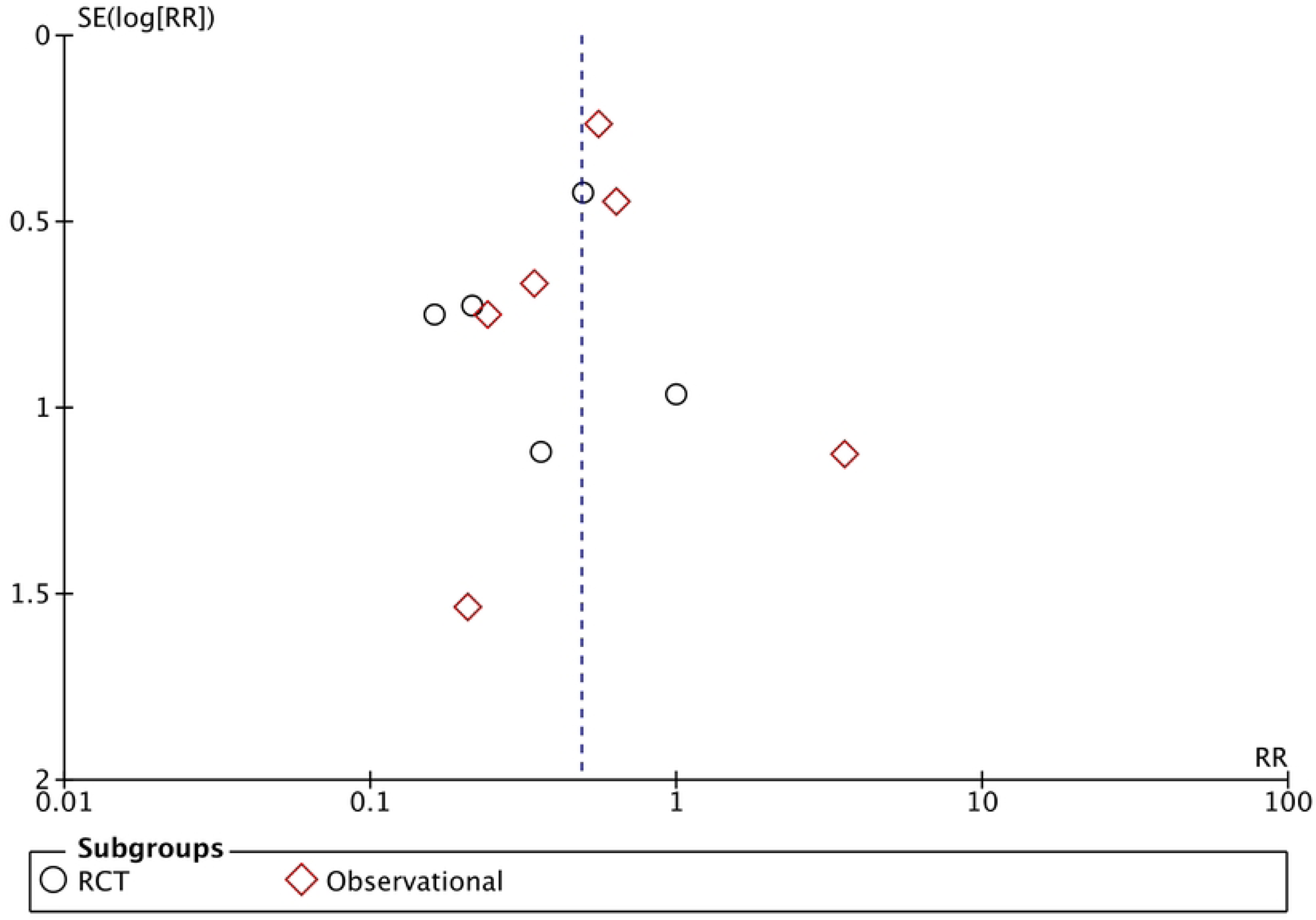

